# Diagnostic Performance of Radiomics in Prediction of Ki-67 Index Status in Non-small Cell Lung Cancer: A Systematic Review and Meta-Analysis

**DOI:** 10.1101/2024.01.11.24301131

**Authors:** Ramin Shahidi, Ehsan Hassannejad, Mansoureh Baradaran, Michail E. Klontzas, Zanyar HajiEsmailPoor, Weelic Chong, Nima Broomand, Mohammadreza Alizadeh, Hamidreza Sadeghsalehi, Navid Mozafari, Soraya Teimoori, Akram Farhadi, Hamed Nouri, Parnian Shobeiri, Houman Sotoudeh

**Author notes:** Corresponding Author: Houman Sotoudeh M.D., Associate Professor of Radiology (P) and Neurology (S); Director of Clinical Research in Neuroradiology Section; Associate Scientist (C), O’Neal Comprehensive Cancer Center; UAB | The University of Alabama at Birmingham, JTN 333, 619 19th St S, Birmingham, AL 35294.

## Abstract

**Background:** Lung cancer is a global health concern, in part due to its high prevalence and invasiveness. The Ki-67 index, indicating cellular proliferation, is pivotal for assessing lung cancer aggressiveness. Radiomics is the inference of quantifiable data features from medical images through algorithms and may offer insights into tumor behavior. Here, we perform a systematic review and meta-analysis to assess the performance of radiomics for predicting Ki-67 status in Non-small Cell Lung Cancer (NSCLC) on CT scan.

**Methods and materials:** A comprehensive search of the current literature was conducted using relevant keywords in PubMed/MEDLINE, Embase, Scopus, and Web of Science databases from inception to November 16, 2023. Original studies discussing the performance of CT-based radiomics for predicting Ki-67 status in NSCLC cohorts were included. The quality assessment involved quality assessment of diagnostic accuracy studies (QUADAS-2) and radiomics quality score (RQS). Quantitative meta-analysis, using R, assessed pooled sensitivity and specificity in NSCLC cohorts.

**Results:** We identified 10 studies that met the inclusion criteria, involving 2279 participants, with 9 of these studies included in quantitative meta-analysis. The overall quality of the included studies was moderate to high based on QUADAS-2 and RQS assessment. The pooled sensitivity and specificity of radiomics-based models for predicting the Ki-67 status of NSCLC training cohorts were 0.78 (95% CI [0.73; 0.83]) and 0.76 (95% CI [0.70; 0.82]), respectively. The pooled sensitivity and specificity of radiomics-based models for predicting the Ki-67 status of NSCLC validation cohorts were 0.79 (95% CI [0.73; 0.84]) and 0.69 (95% CI [0.61; 0.76]), respectively. Substantial heterogeneity was noted in the pooled sensitivity and specificity of training cohorts and the pooled specificity of validation cohorts (I^2^ > 40%). It was identified that utilizing ITK-SNAP as a segmentation software contributed to a significantly higher pooled sensitivity.

**Conclusion:** This meta-analysis indicates promising diagnostic accuracy of radiomics in predicting Ki-67 in NSCLC. The study underscores radiomics’ potential in personalized lung cancer management, advocating for prospective studies with standardized methodologies and larger samples.

## 1. Introduction

Lung cancer is a serious global health concern since it is one of the most malignant tumors, in part due to its late detection and high mortality (1). Improving diagnostic accuracy and prognostic assessment is critical to enhance patient outcomes. The Ki-67 index is a biochemical marker associated with cellular proliferation in lung malignancies that is critical in evaluating tumor aggressiveness and the effectiveness of treatment regimens, a main topic of interest in the field (2). Although not part of the diagnostic criteria, the Ki-67 index is important in personalized medicine, allowing individualized therapy approaches based on individual tumor features (3).

Radiomics, a novel medical imaging method, may improve lung cancer diagnosis by extracting quantitative data from common imaging modalities such as computed tomography (CT) scans and magnetic resonance imaging (MRIs) (4). This approach provides novel insights into tumor heterogeneity and behavior through the automated extraction and analysis of detailed tumor features such as texture and shape (5). Building upon non-invasive and repeatable imaging modalities, radiomics provides insights into lung cancer progression and therapy response (6).

The increased interest in using radiomics to predict Ki-67 index status in lung tumors highlights the technology’s potential to improve cancer diagnosis and treatment planning. Radiomics utilizes the latest imaging techniques, such as CT and positron emission tomography (PET) scans, to estimate the Ki-67 index non-invasively, linking radiomics features with tumor proliferation rates (7). This strategy can potentially transform lung cancer treatment by encouraging the development of personalized therapies based on tumor features.

In light of this, this meta-analysis aims to assess radiomics’ diagnostic accuracy in predicting the Ki-67 index in Non-small Cell Lung Cancer (NSCLC). It aims to synthesize existing research to guide future therapeutic applications, thereby filling a knowledge gap. By giving a complete overview of radiomics’ capability to predict the Ki-67 index, this work has the potential to have a substantial impact on the approach to personalized lung cancer management.

## 2. Methods and Materials

This comprehensive review followed the standards set forth in the Preferred Reporting Items for Systematic Reviews and Meta-analysis (PRISMA) statement (8). The research plan was officially recorded in the International Prospective Register of Systematic Reviews (PROSPERO) under the identifier CRD42023487733.

### 2.1 Search Strategy

A thorough search of existing literature was conducted to locate observational studies investigating the prediction potential of CT scan-derived radiomics in identifying the Ki-67 index status in lung cancers. This involved systematically searching the Scopus, Embase, Web of Science and PubMed/MEDLINE databases until November 16, 2023, using specific keywords like “Radiomic*”, “Lung”, “Pulmonary”, “Ki-67”, “Ki67”, “MIB-1”, and “MIB1”. Unique search strategies were crafted for each database, employing specific keywords and suitable Boolean operators (such as OR/AND). No restrictions were placed on publication dates, study designs, languages, or publication countries to ensure a comprehensive review of the existing literature.

### 2.2 Inclusion and Exclusion Criteria

This research encompassed various observational inquiries, consisting of retrospective and prospective cross-sectional and cohort studies. These investigations delved into how effective CT scan-based radiomics are in predicting the Ki-67 index status in cases of lung cancers. We included studies with the following criteria: (1) Age ≥ 18; (2) Individuals diagnosed with lung cancers, irrespective of the specific histological subtype; (3) Studies utilizing CT scan-based radiomics analysis to predict Ki-67 index status, with reported diagnostic performance metrics like sensitivity and specificity and outcomes related to distinct Ki-67 index categories (e.g., low, high).

Exclusion criteria encompassed: (1) Other study types (2) Studies involving pediatric populations (age below 18 years); (3) Studies exclusively focused on patients with tumors other than lung primary tumors; (4) Studies not utilizing radiomics analysis for predicting Ki-67 index status; (5) Studies employing imaging modalities other than CT scan for radiomics analysis; (6) Studies lacking relevant diagnostic performance metrics or outcomes related to Ki-67 index categories.

### 2.3 Study Selection Process

In the first stage, we imported the retrieved records into the EndNote 20 software (Clarivate Analytics, Philadelphia, PA, USA), and subsequently duplicate entries were removed. In an initial screening, two reviewers (S.T and M.B) evaluated the eligibility of studies based only on their titles and abstracts, working independently. The reviewers performed a subsequent round of assessment for the selected articles, appraising their full texts according to pre-established criteria for inclusion and exclusion. To resolve any discrepancies, a third reviewer (R.S) was consulted to achieve an agreement.

### 2.4 Data Extraction

Two independent investigators (N.M and E.H) meticulously examined the included studies, and utilized a predefined Microsoft Excel worksheet to extract the following information: name of the first author, year of publication, country of study, study design, the number and demographic data of patients, CT scan manufacturer, segmentation type, the modeling classifier, reference standard (ground-truth), radiomics feature extraction and selection details (such as the software, extracted features, selected features, feature extraction and selection methods), and diagnostic performance metrics of radiomics-based machine learning model (including sensitivity and specificity). A third reviewer (R.S) conducted a final check on the extracted data. In instances where the values for True Positive (TP), True Negative (TN), False Positive (FP), and False Negative (FN) are not explicitly provided, we reconstructed these parameters utilizing the reported sensitivity and specificity, along with the number of patients with high and low Ki-67 expression levels. We rounded the resulting values to the nearest whole numbers, particularly if they were in decimal form. Consequently, a marginal variance between the sensitivity and specificity values presented in our article and those in the referenced articles may arise due to the applied rounding procedure. Whenever a study provided only a receiver operating characteristic (ROC) curve without sensitivity and specificity values in the text, we applied the top-left method for extracting sensitivity and specificity.

### 2.5 Quality Assessment

Two independent researchers (HR.S and S.T, both with 2 years of radiomics research experience) evaluated the methodical rigorousness of the studies included in this research using two assessment instruments: the Quality Assessment of Diagnostic Accuracy Studies (QUADAS-2) (9) and the Radiomics Quality Score (RQS) (10). A third reviewer (H.S, with 5 years of radiomics research experience), also working independently and blinded to the assessments of the other reviewers, rechecked this stage.

The QUADAS-2 instrument comprises four key areas: index test, patient selection, reference standard, and flow and timing. Every domain undergoes meticulous scrutiny to identify possible biases, while assessing the relevance of index tests, patient selection, and reference standards. This evaluation categorizes each study’s bias risk as low, high, or indeterminate. Additionally, the tool offers guiding questions to assist in discerning potential bias. Additionally, the RQS, a tool tailored for assessing the quality of radiomics studies, comprises sixteen specific methodological items. These items collectively contribute to a maximum score of 36, offering a consensus-based evaluation of radiomics study quality.

### 2.6 Quantitative Meta-Analysis

#### 2.6.1 Software

All calculations, visual representations, and additional analysis for meta-analyses with considerable heterogeneity were conducted using R software version 4.3.2 along with the ‘dmetar,’ ‘meta,’ and ‘tidyverse’ packages. (11) (R Core Team (2021). R: A language and environment for statistical computing. R Foundation for Statistical Computing, Vienna, Austria. https://www.R-project.org/.).

#### 2.6.2 Statistical Analysis

The chosen level of significance for statistical analysis stood at p-values below 0.05. The effect size was quantified by evaluating the AI model’s performance in distinguishing between high and low Ki-67 expression groups. The analytical model comprised both fixed and random effects concurrently. Heterogeneity was assessed using the I^2^ index, where an I^2^ value < 40% suggested insignificant inconsistency across studies, prompting the use of a fixed-effects model for meta-analysis. Conversely, if I^2^ estimates exceeded 40%, the analysis employed a random-effects approach.

#### 2.6.3 Sensitivity analysis and subgroup analysis

We conducted a sensitivity analysis on results that showed significant heterogeneity (I^2^>40%) in order to enhance our understanding of possible sources of heterogeneity. Within this analysis, we methodically omitted one study at a time and re-evaluated the effect size, employing a Leave-One-Out method, aiming to refine our understanding of heterogeneity sources. In addition, to explore potential sources of heterogeneity, we performed subgroup analyses based on tumor type (Adenocarcinoma vs. non-specific NSCLC), sample size (more than 200 vs. less than 200), cut-off for Ki-67 positivity (more than 20% vs. less than 20%), segmentation software (ITK-SNAP vs. other software), study design (multicenter study vs. single center), and RQS grade (above or equal to average vs. below than average). This comprehensive approach allowed for a more assessment of factors contributing to the observed heterogeneity.

## Results

### Characteristics of Included Studies

A comprehensive database search initially yielded ninety-six records. After removing duplicate records, the remaining 52 records underwent title and abstract screening. Out of these, 21 records underwent full-text screening, and ultimately, ten articles encompassing 2279 patients were selected for qualitative analysis, and after a meticulous screening process, nine of them were included in the quantitative analysis. (Fig. 1). All studies were designed retrospectively and conducted in China. Table 1 offers a comprehensive overview of studies focusing on radiomics for predicting Ki-67 expression in NSCLC.

**Fig 1.**
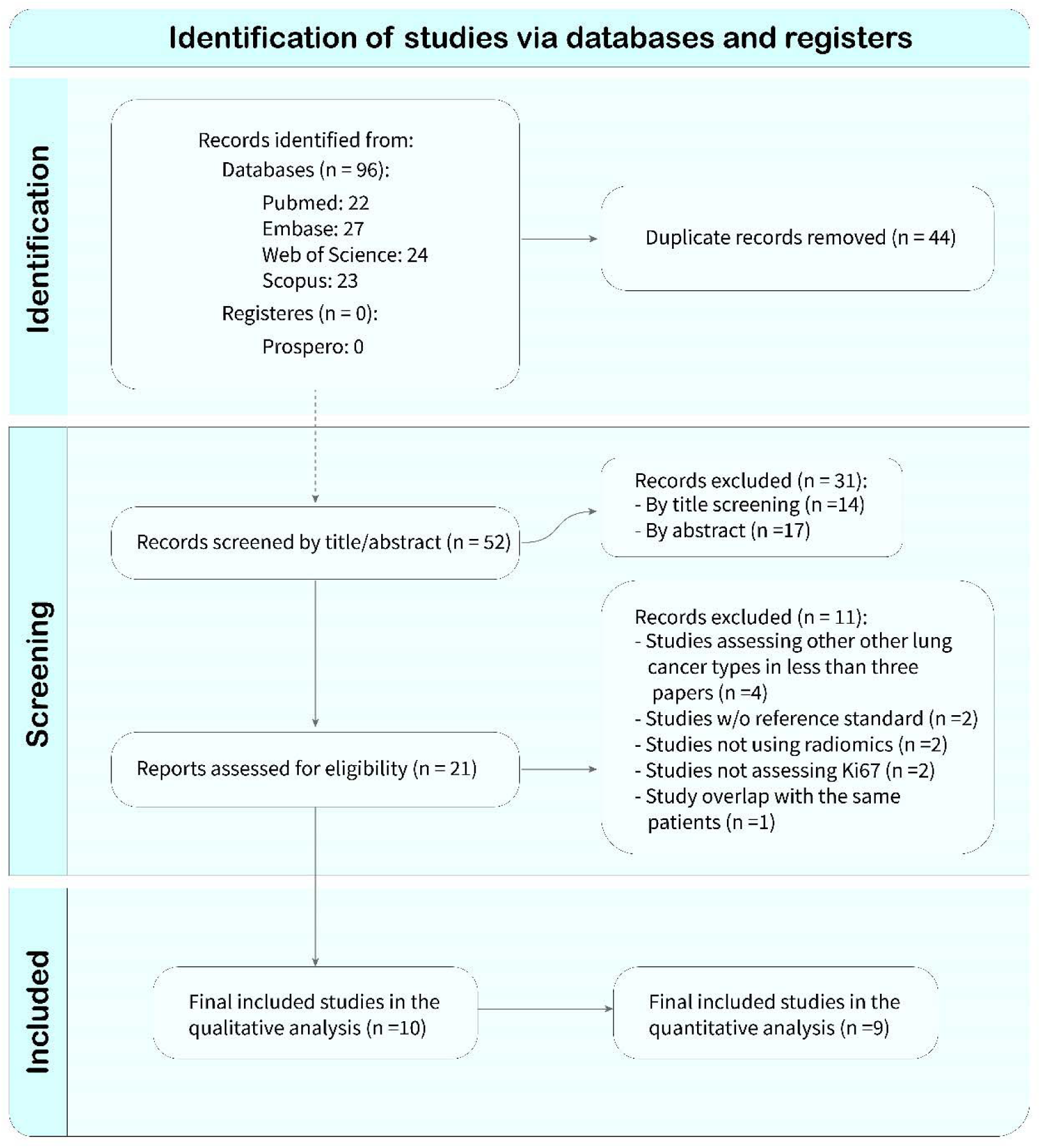
PRISMA Flowchart illustrating the process of selecting eligible studies.

**Table 1.**
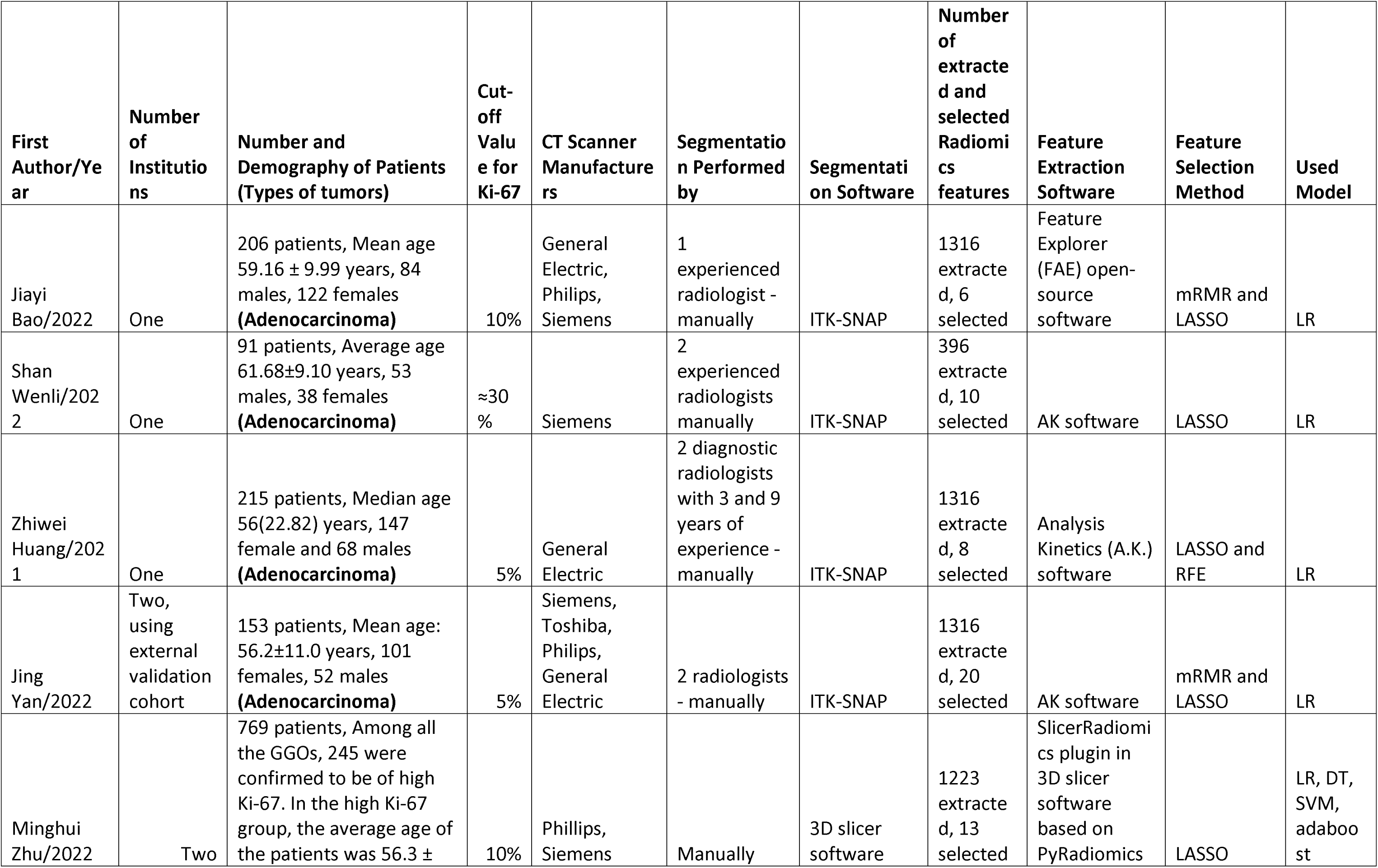
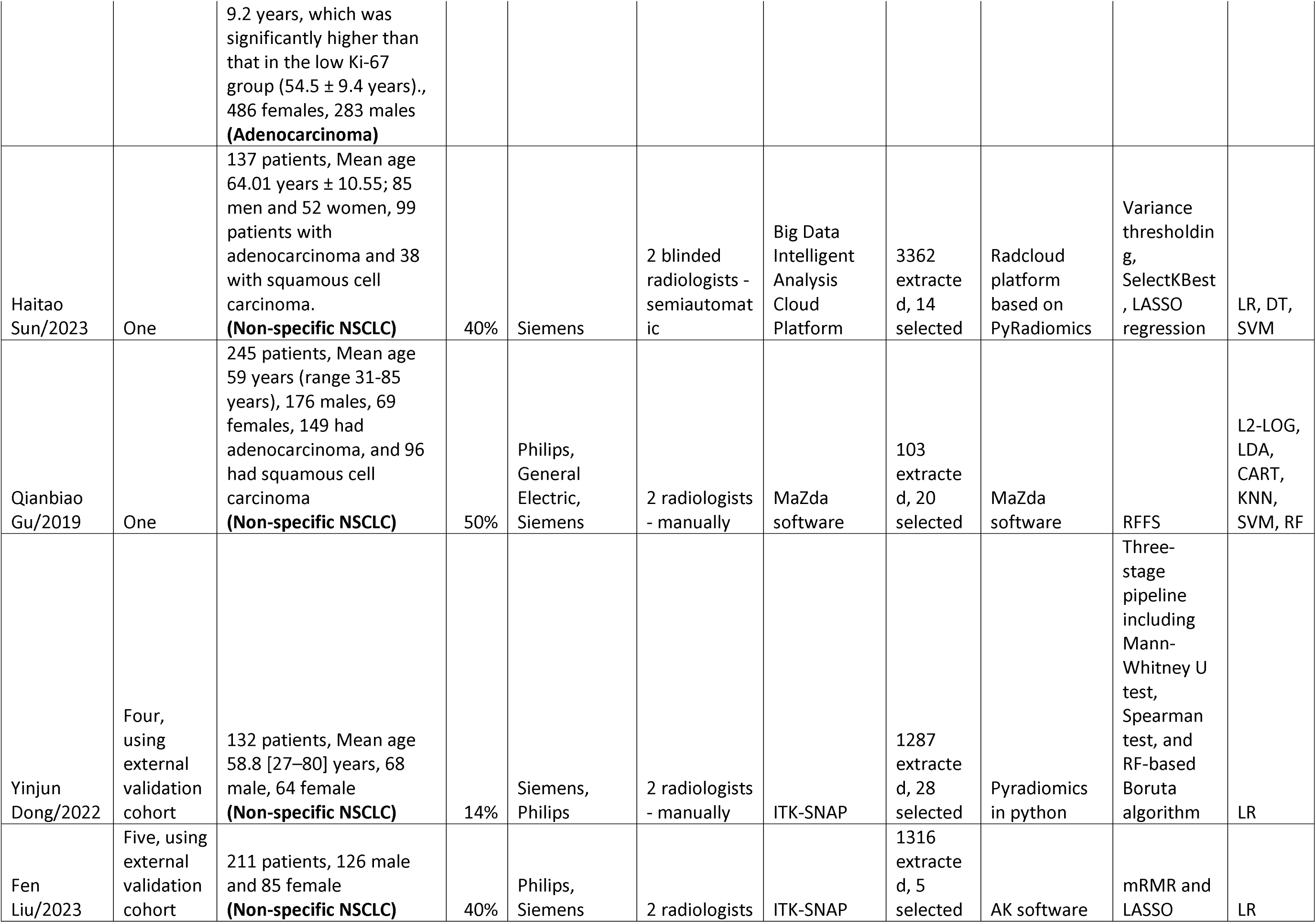
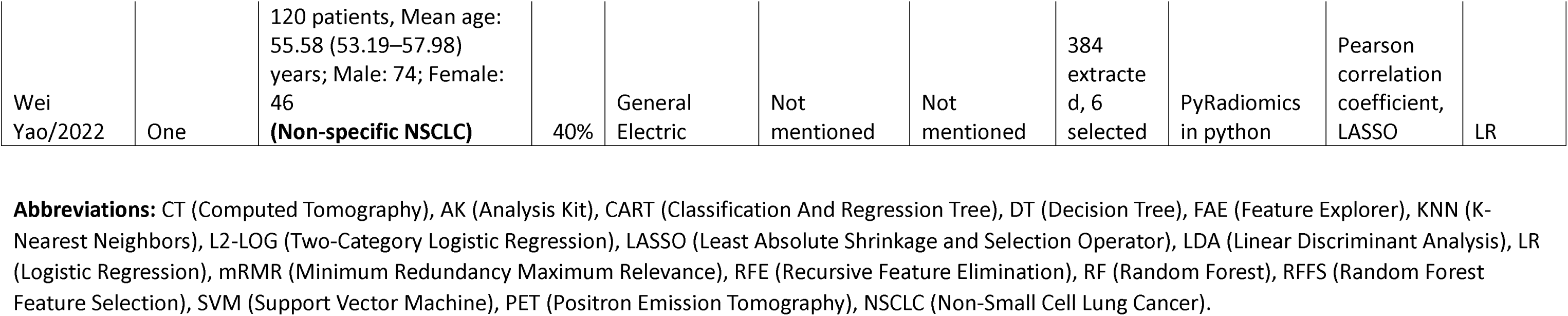
Overview of Studies on Radiomics for Predicting Ki67 Expression in Lung Cancers.

We observed a spectrum of cut-off values for Ki-67 across studies, encompassing values of 5% (12, 13), 10% (14, 15), 14% (16), 30% (17), 40% (18–20), and 50% (21). Most studies utilized immunohistochemical analysis to assess the Ki-67 index in samples obtained from surgeries or biopsies (12–15, 18, 19, 21).

Image segmentation was mainly carried out manually by one or two skilled radiologists (12–18, 21). However, in a particular study, two blinded radiologists were involved in performing semi-automatic segmentation (19).

The predominant segmentation tool employed across the majority of studies was ITK-SNAP (60%) (12–14, 16–18). A single study utilized MaZda software (21), another study leveraged 3D Slicer (15). Additionally, a distinct study utilized the specialized Big Data Intelligent Analysis Cloud Platform (19).

Different software tools were used in extracting radiomic features across studies. GE Healthcare’s Analysis Kit (AK) software emerged as a common choice, employed in multiple studies for this purpose (12, 13, 17, 18). Additionally, Feature Explorer (FAE) open-source software (14), the 3D Slicer PyRadiomics plugin (15), the python-based PyRadiomics implementation (16, 20), the Radcloud platform (19), and MaZda software (21) were among the tools utilized.

Various methods for selecting radiomics features were utilized in the included papers, with a majority incorporating Least Absolute Shrinkage and Selection Operator (LASSO) in 80% (12–15, 17–20) of the studies and minimum redundancy maximum relevant (mRMR) in 30% (12, 14, 18) of them. Additionally, other methods were employed, such as Recursive Feature Elimination (RFE) (13), variance thresholding (19), SelectKBest (19), Random Forest Feature Selection (RFFS) (21), Pearson correlation coefficient (20), and a combination of Mann-Whitney U test, Spearman test, RF-based Boruta algorithm (16). These techniques were used by the authors to select features, with the number selected ranging from 5 to 28 per study.

Most studies in our review demonstrated a strong preference for logistic regression models in their radiomic analysis, with some incorporating additional methodologies for enhanced robustness and precision (12–14, 16–18, 20). In contrast, Zhu et al. and Sun et al. diversified their approach by integrating logistic regression with decision trees, support vector machines (SVM), and adaboost (15, 19). Notably, Gu et al. used a variety of modeling approaches, including two-category logistic regression (L2-LOG), linear discriminant analysis (LDA), classification and regression trees (CART), K-nearest neighbors (KNN) clustering, radial SVM, and random forest (RF) (21).

### Quality Assessment

The quality assessment of the included studies was performed using both QUADAS-2 and the RQS. The QUADAS-2 risk of bias assessment revealed that the majority of studies exhibited a low risk in patient selection, index test, and flow and timing, albeit with some uncertainties in the reference standard. In terms of QUADAS-2 applicability, most studies demonstrated low concerns in patient selection, index test, and reference standard. (Fig. 2 and 3) The RQS assessment displayed a wide range of scores, from 1 (2.78%) to 19 (52.78%), indicating a significant variation in the quality of the studies (Fig. 4). Dong et al. (16) achieved the highest RQS of 19 (52.78%), while Gu et al. (21) scored the lowest with 1 (2.78%). These results underscore the importance of rigorous and standardized reporting in radiomics studies. (Table. 1 and 2 in Supplementary Materials)

**Fig 2.**
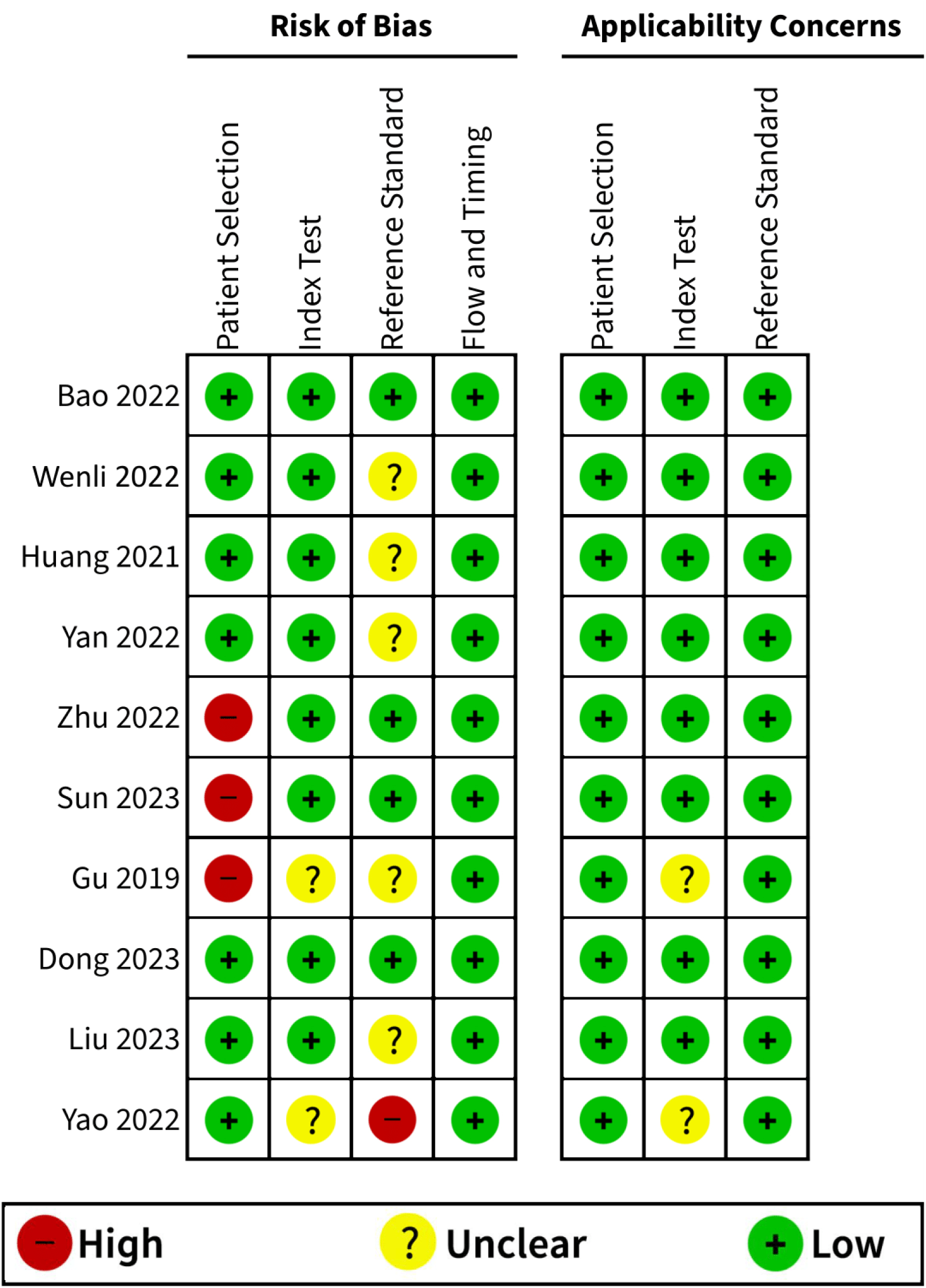
Critical appraisal based on the QUADAS-2 tool - part 1.

**Fig 3.**
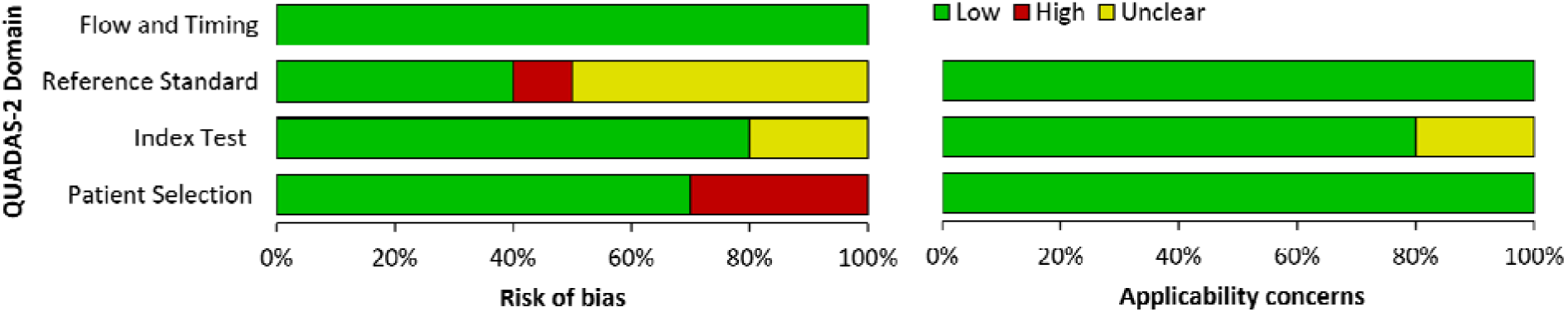
Critical appraisal based on the QUADAS-2 tool - part 2.

**Fig 4.**
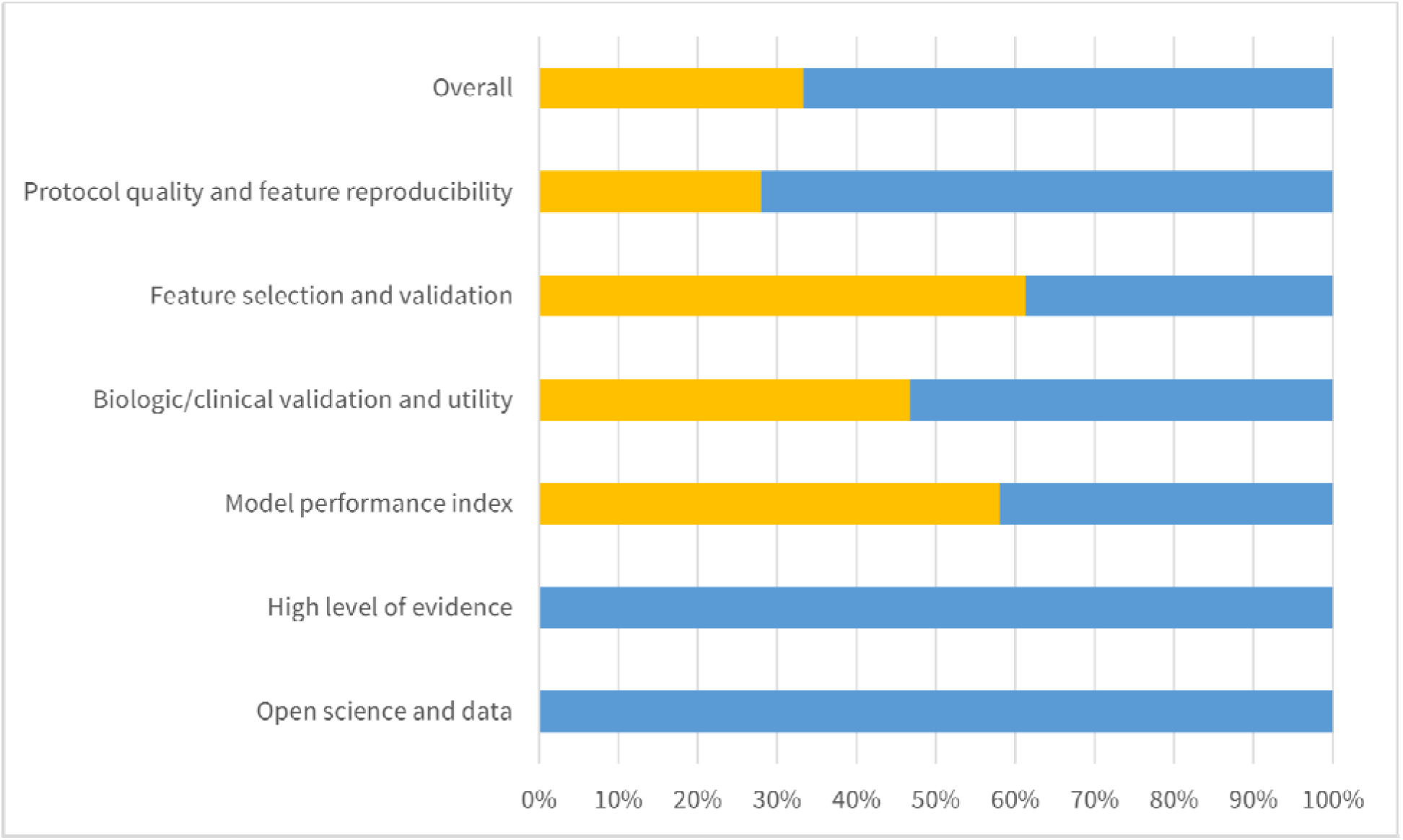
Critical appraisal based on the RQS tool.

The overall adherence to the six key domains of the RQS stood at 33.3%, indicating a general adherence to recommended practices across the included studies. However, certain areas showed weaknesses, particularly in the domain of protocol quality and stability in image and segmentation, with an adherence rate of only 28%. Feature selection and validation had the highest adherence rate at 61.3%, followed by biologic/clinical validation and utility at 46.7%. The model performance index domain scored 58% in adherence, while high-level evidence and open science and data domains had the lowest adherence rates, both at 0%.

### Meta-Analysis

An overview of pooled sensitivity and specificity of training and validation cohorts of NSCLC have been provided.

### Training Cohorts

Data from nine studies were used for assessing the sensitivity and specificity of radiomics-based models to predict Ki-67 status in training cohorts of patients with NSCLC. The pooled sensitivity for this task was 0.78 (95% CI [0.73; 0.83], I² = 47%, p = 0.06), while the pooled specificity stood at 0.76 (95% CI [0.70; 0.82], I² = 72%, p L 0.01). (Fig. 5A and Fig. 5B) Given the considerably high I² level in pooled sensitivity and specificity, a sensitivity analysis was conducted, revealing that the study by Sun et al. (19) had the most influence on heterogeneity reduction when removed. (Fig. 1S) Also, this study acted as an outlier in pooled specificity, and upon its removal, the heterogeneity of pooled specificity significantly reduced to 0.7465 (95% CI [0.6885; 0.7969], I² = 60.6%, p-value = 0.013).

**Fig 5.**
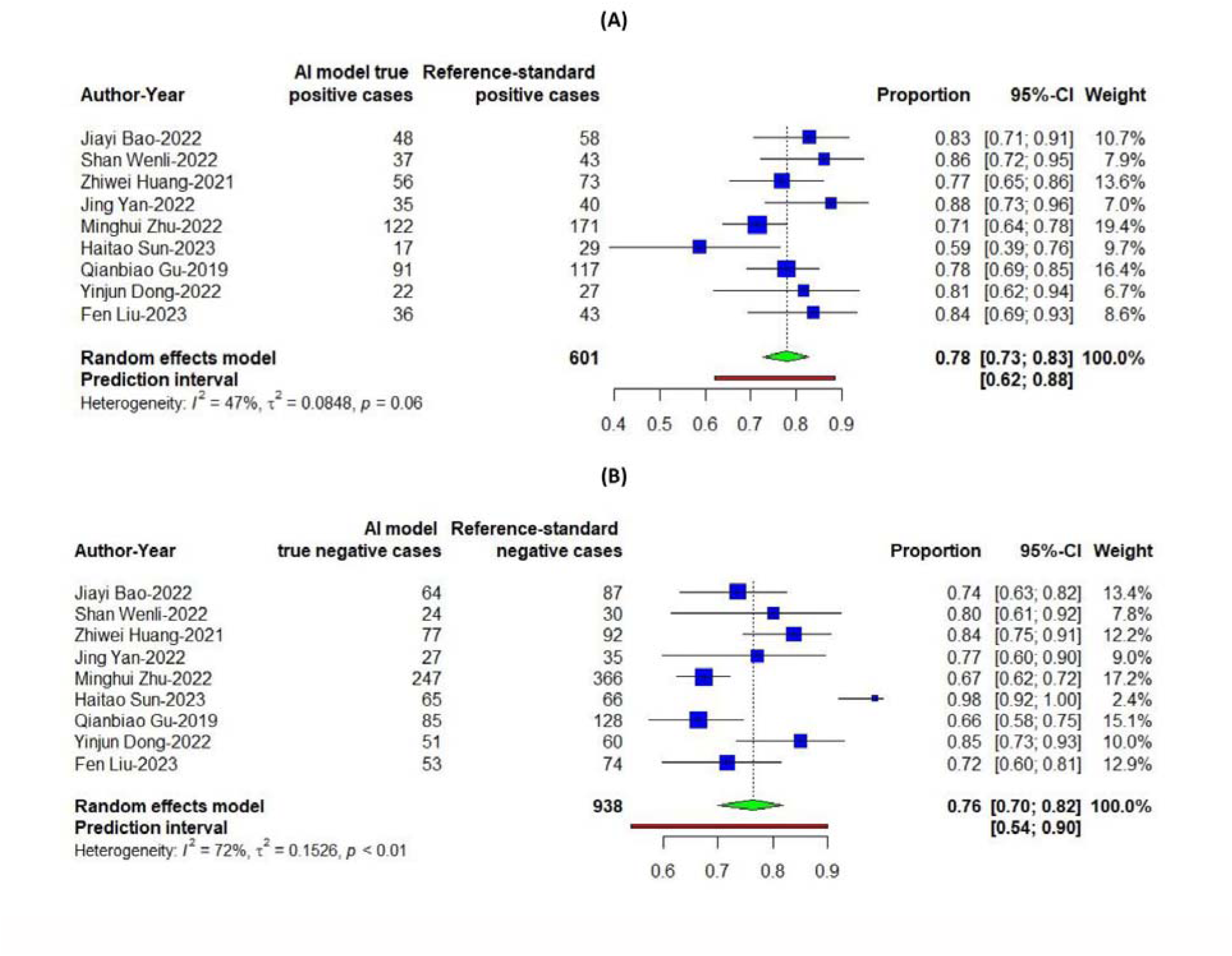
Pooled estimates of training cohorts (A) Sensitivity, (B) Specificity.

### Validation Cohorts

Data from eight studies were utilized to assess the performance of radiomics-based models in predicting Ki-67 status in validation cohorts of patients with NSCLC. The pooled sensitivity, calculated at 0.79 (95% CI [0.73; 0.84]), exhibited low heterogeneity (I² = 0, p = 0.65). Conversely, the pooled specificity stood at 0.69 (95% CI [0.61; 0.76]) with a higher level of heterogeneity (I² = 44%, p = 0.09) (Fig. 6A and Fig 6B). A sensitivity analysis was conducted due to the notable specificity’s heterogeneity, highlighting the study by Zhu et al. (15) as the primary contributor to reducing heterogeneity when omitted. (Fig. 1S)

**Fig 6.**
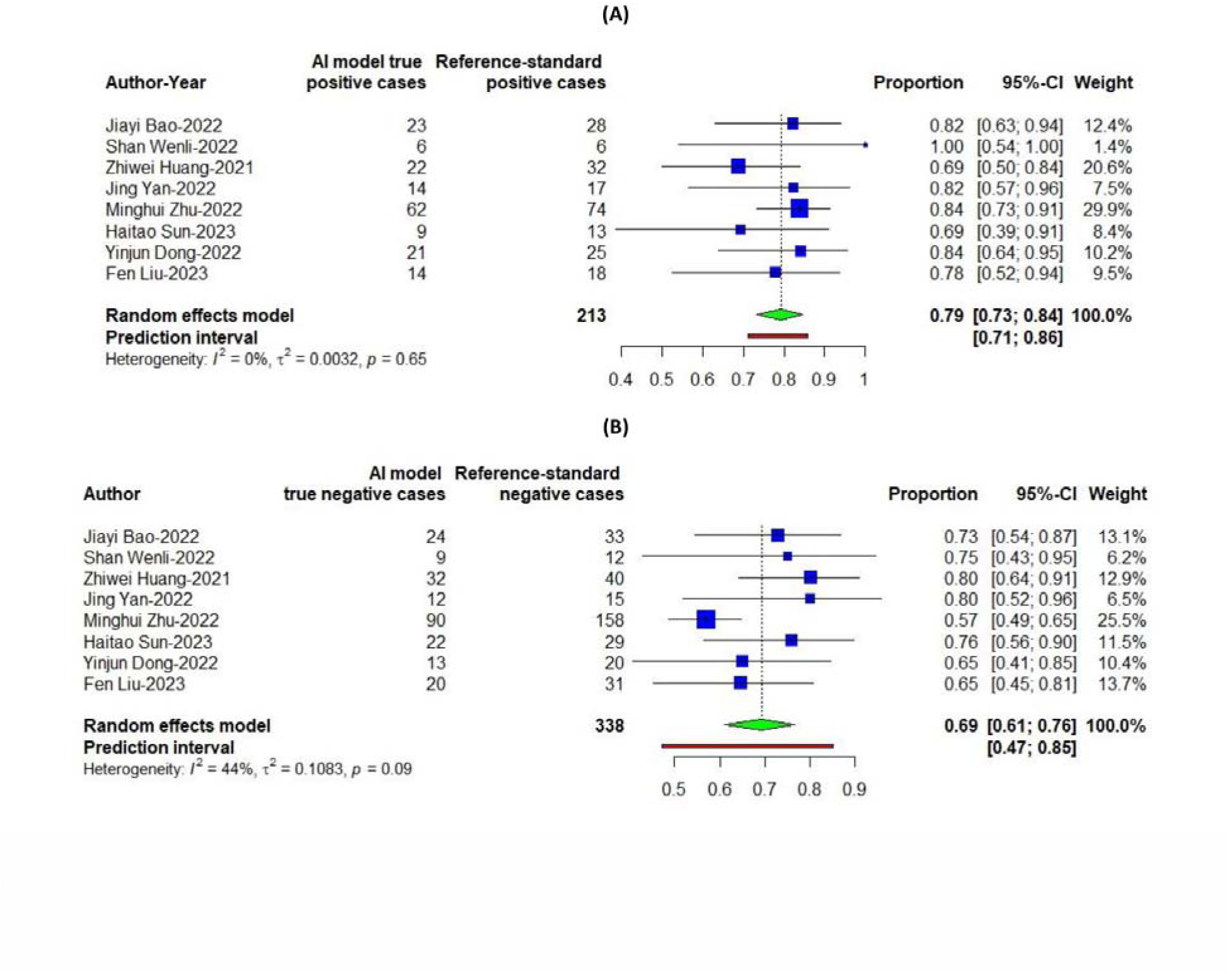
Pooled estimates of Validation cohorts (A) Sensitivity – Validation cohort, (B) Specificity – Validation cohort.

### Subgroup analysis

According to the subgroup analysis in training cohorts, studies which used ITK-SNAP as a segmentation software had a higher pooled sensitivity (82% vs. 71%) (p-value=0.02) and lower pooled specificity (78% vs. 84%) (p-value=0.72, not significant). In addition, in studies with sample sizes smaller than 200, pooled sensitivity (80% vs. 77%) (p-value=0.69, not significant) and pooled specificity (85% vs. 72%) (p-value=0.048) was higher than those studies that included more than 200 patients. Also, our subgroup analysis in validation cohorts revealed that studies with multicenter design had a higher pooled sensitivity (83% vs. 74%) (p-value=0.15, not significant) and lower pooled specificity (61% vs. 76%) (p-value=0.013). The results of the subgroup analysis are summarized in Table 2 and 3.

**Table 2.** Subgroup analysis for Training cohorts.

|  | Number of studies | Sensitivity |  |  | Specificity |  |  |
| --- | --- | --- | --- | --- | --- | --- | --- |
|  |  | Pooled sensitivity | I<sup>2</sup> | P-value (between study differences) | Pooled sensitivity | I<sup>2</sup> | P-value (between study differences) |
| Type of cancer |  |  |  |  |  |  |  |
| Adenocarcinoma | 5 | 0.7952 [0.7223; 0.8529] | 52.50% | 0.5646 | 0.7542 [0.6790; 0.8165] | 63.90% | 0.4514 |
| Non-specific NSCLC | 4 | 0.7613 [0.6508; 0.8451] | 54.30% |  | 0.8342 [0.5947; 0.9452] | 82.30% |  |
| Sample size |  |  |  |  |  |  |  |
| More than 200 | 5 | 0.7686 [0.7178; 0.8126] | 20.80% | 0.6895 | 0.7201 [0.6559; 0.7765] | 61.20% | 0.048 |
| Less than 200 | 4 | 0.7964 [0.6469; 0.8931] | 69.10% |  | 0.8508 [0.7333; 0.9221] | 61.50% |  |
| Cut-off for Ki-67 Positivity |  |  |  |  |  |  |  |
| More than 20 | 4 | 0.7759 [0.6547; 0.8634] | 63.60% | 0.9088 | 0.8202 [0.5739; 0.9392] | 77.00% | 0.6302 |
| Less than 20 | 5 | 0.7829 [0.7141; 0.8389] | 40.00% |  | 0.7688 [0.6889; 0.8332] | 73.40% |  |
| Segmentation software |  |  |  |  |  |  |  |
| ITK-SNAP | 6 | 0.8208 [0.7711; 0.8616] | 0.00% | 0.0214 | 0.7816 [0.7268; 0.8280] | 19.50% | 0.7199 |
| Other softwares | 3 | 0.7149 [0.6244; 0.7910] | 55.00% |  | 0.8389 [0.4064; 0.9754] | 82.90% |  |
| Multicenter study? |  |  |  |  |  |  |  |
| Yes | 4 | 0.7979 [0.7009; 0.8692] | 54.40% | 0.6569 | 0.7423 [0.6533; 0.8150] | 62.70% | 0.3846 |
| No | 5 | 0.7727 [0.6941; 0.8358] | 51.40% |  | 0.8071 [0.6692; 0.8964] | 78.20% |  |
| RQS grade |  |  |  |  |  |  |  |
| Above or equal to average | 5 | 0.7944 [0.6923; 0.8691] | 56.20% | 0.5489 | 0.8098 [0.6997; 0.8861] | 68.50% | 0.2442 |
| Below average | 4 | 0.7620 [0.7047; 0.8111] | 39.00% |  | 0.7317 [0.6360; 0.8098] | 71.40% |  |

| Sensitivity | Specificity |
| --- | --- |

**Table 3.** Subgroup analysis for validation cohorts.

|  | Number of studies | Pooled sensitivity | I^2 | P-value (between study differences) | Pooled sensitivity | I^2 | P-value (between study differences) |
| --- | --- | --- | --- | --- | --- | --- | --- |
| Type of cancer |  |  |  |  |  |  |  |
| Adenocarcinoma | 5 | 0.8000 [0.7157; 0.8641] | 0.00% | 0.7695 | 0.7100 [0.5885; 0.8073] | 63.10% | 0.7423 |
| Non-specific NSCLC | 3 | 0.7804 [0.6509; 0.8713] | 0.00% |  | 0.6846 [0.5742; 0.7775] | 0.00% |  |
| Sample size |  |  |  |  |  |  |  |
| More than 200 | 4 | 0.7889 [0.7025; 0.8553] | 5.40% | 0.8142 | 0.6750 [0.5565; 0.7746] | 64.50% | 0.4427 |
| Less than 200 | 4 | 0.8043 [0.6819; 0.8874] | 0.00% |  | 0.7336 [0.6221; 0.8216] | 0.00% |  |
| Cut-off for Ki-67 Positivity |  |  |  |  |  |  |  |
| More than 20 | 3 | 0.7606 [0.5933; 0.8737] | 0.00% | 0.5894 | 0.7054 [0.5893; 0.7998] | 0.00% | 0.8735 |
| Less than 20 | 5 | 0.8019 [0.7288; 0.8590] | 0.00% |  | 0.6932 [0.5789; 0.7878] | 60.80% |  |
| Segmentation software |  |  |  |  |  |  |  |
| ITK-SNAP | 6 | 0.7829 [0.7003; 0.8477] | 0.00% | 0.8198 | 0.7237 [0.6457; 0.7900] | 0.00% | 0.4555 |
| Other softwares | 2 | 0.7996 [0.6525; 0.8945] | 33.40% |  | 0.6499 [0.4481; 0.8094] | 71.40% |  |
| Multicenter study? |  |  |  |  |  |  |  |
| Yes | 4 | 0.8275 [0.7537; 0.8826] | 0.00% | 0.1506 | 0.6133 [0.5287; 0.6916] | 11.40% | 0.0131 |
| No | 4 | 0.7434 [0.6328; 0.8297] | 0.00% |  | 0.7619 [0.6748; 0.8314] | 0.00% |  |
| RQS grade |  |  |  |  |  |  |  |
| Above or equal to average | 5 | 0.8003 [0.7028; 0.8716] | 0.00% | 0.8471 | 0.7017 [0.6152; 0.7759] | 0.00% | 0.9299 |
| Below average | 3 | 0.7873 [0.6683; 0.8718] | 35.60% |  | 0.7102 [0.5184; 0.8480] | 77.20% |  |

## Discussion

Ki-67, a cell proliferation-associated antigen, is predominantly found in the nucleus and plays a critical role in cell growth by correlating with mitotic activity and the cell cycle. It is particularly significant in marking cells in the growth phase and is used to determine tumor cell proportions, linking with rapid tumor growth and a worse prognosis. Specifically in lung cancer, cells expressing Ki-67 exhibit heightened proliferation, greater invasiveness, and a higher likelihood of lymph node metastasis (22). Its higher expression might also be an indicator of shortened progression-free survival (PFS) time (23). In addition, it yields prognostic importance in lung adenocarcinoma subtypes, suggests its potential as a marker in lung neuroendocrine tumors, and identifies its clinical significance in advanced NSCLC, particularly in predicting chemotherapy efficacy (23, 24).

Radiomics, initially developed for oncologic imaging to aid personalized medicine with imaging biomarkers, is adaptable for all pathologies and can analyze any digital images (25). This quantitative approach has been successfully employed in diagnosing, differentiating, grading, post-treatment evaluation, and survival prediction of different types of tumors (26, 27).

In the present review, we explored the applicability of radiomics approaches in predicting the Ki-67 proliferation index in NSCLC. Our meta-analysis revealed promising observations regarding the sensitivity and specificity of these models in predicting the Ki-67 status of NSCLC. The pooled sensitivity and specificity rates were generally high, but with moderate to high heterogeneity among studies.

In further exploration through subgroup analysis in training cohorts, we observed that studies employing ITK-SNAP as a segmentation software exhibited a significantly higher pooled sensitivity and a nonsignificant decrease in pooled specificity. The heightened sensitivity associated with the use of ITK-SNAP in segmentation software may be due to its relatively fast segmentation time, intuitive usability, good 3D visualization, easy handling of segmentation outputs, and multi-platform support (28).

While most of the studies employed logistic regression models, studies by Zhu et al., Sun et al., and Gu et al. integrated these models with a range of other machine learning approaches such as SVM, DT, L2-LOG, LDA, CART, KNN, RF, and Adaboost (15, 19, 21). Yet, these integrations did not lead to a better performance, i.e. the machine learning models have lower sensitivity and specificity. Fast training, versatility in handling discrete and continuous variables, and strong predictive performance in cancer patient outcomes like tumor mutation burden, survival time, and treatment response are the main strengths of logistic regression models (29–32).

However, logistic regression models may face significant challenges, including reduced interpretability with a large number of features, susceptibility to multicollinearity and overfitting, suboptimal performance in complex nonlinear scenarios, difficulties in integrating diverse data types, and confounding effects from tumor volume that can compromise both model performance and interpretability (33, 34). Yet, through our analyses, these models appeared to perform the best when predicting Ki-67 expression in lung cancers.

Beyond Ki-67, key biomarkers in cancer radiomics include epidermal growth factor (EGFR) and isocitrate dehydrogenase (IDH). EGFR, a crucial biomarker in lung cancer, particularly NSCLC, is linked to cancer growth and progression. It has also been the most frequently analyzed biomarker in cancer radiomics studies. IDH, third in analysis frequency, impacts metabolic regulation and cancer progression. Future studies may explore integrating these biomarkers into machine-learning models for enhanced insights (35, 36).

## Limitations

Despite the promising findings of these explorations, there were also limitations in our study.

Firstly, all of the included studies in the meta-analysis utilized a retrospective design. This approach has inherent limitations, as it involves analyzing pre-existing data, which can lead to incomplete or missing information. It raises concerns about selection and recall biases, potentially jeopardizing the accuracy and reliability of the findings (37, 38).

Moreover, the lack of a standardized cutoff value for histology-based Ki-67 expression in lung cancer could hinder the applicability of the radiomic models in clinical settings, and further evidence is needed for histology-based and radiomics-based thresholds for the Ki-67 index in lung cancer (18, 19). Future studies should incorporate prospective designs, larger and more diverse samples with external validation, standardized methodologies for data collection and analysis to enhance the reliability, applicability, and precision of radiomics models.

## Conclusion

This meta-analysis assessed the diagnostic accuracy of CT scan-based radiomics in predicting the Ki-67 index in NSCLC. Analyzing ten retrospective studies with 2279 patients primarily from China, the results indicate promising sensitivity and specificity in predicting Ki-67 status in NSCLC cohorts. However, heterogeneity in pooled sensitivity and specificity warrants cautious interpretation. Despite methodological rigor, limitations include retrospective designs and lack of standardized Ki-67 cutoff values. The study underscores radiomics’ potential in personalized lung cancer management, emphasizing the need for prospective studies, larger samples, and standardized methodologies for enhanced reliability. Integrating other biomarkers like EGFR and IDH may further improve predictive capabilities in lung cancer diagnosis and prognosis.

## Supporting information

Supplementary material - Figure 1 - Heterogenity

Supplementary material - Keywords of Radiomic-ki67

Supplementary material - RQS_table_1

Supplementary material - RQS_table_2

Supplementary material - Search Queries of Radiomic-ki67

## Data Availability

All data produced in the present work are contained in the manuscript

## Declaration of Generative AI and AI-assisted Technologies in the Writing

During the preparation of this work, the authors used ChatGPT 3.5 by OpenAI to improve paper readability. After using this service, the authors reviewed and edited the content as needed and took full responsibility for the publication’s content.

