## Supplementary material - Figure 1 - Heterogenity for "Diagnostic Performance of Radiomics in Prediction of Ki-67 Index Status in Non-small Cell Lung Cancer: A Systematic Review and Meta-Analysis"

**(A)**

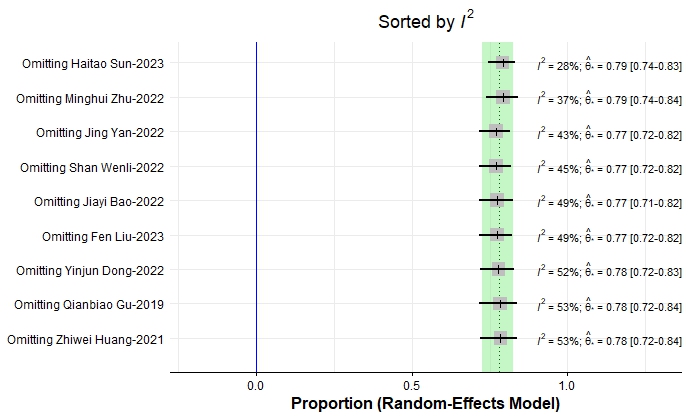

**(B)**

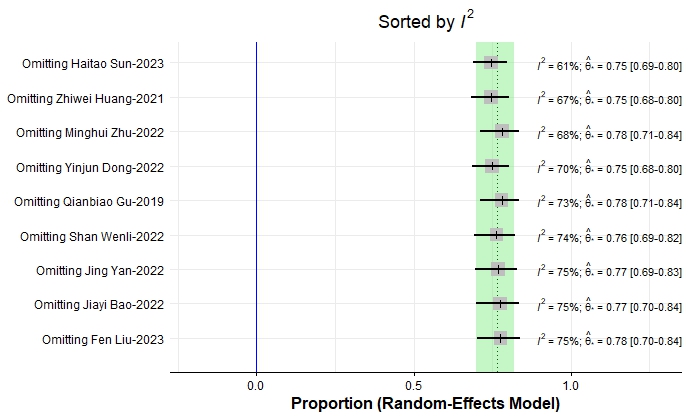

**(C)**

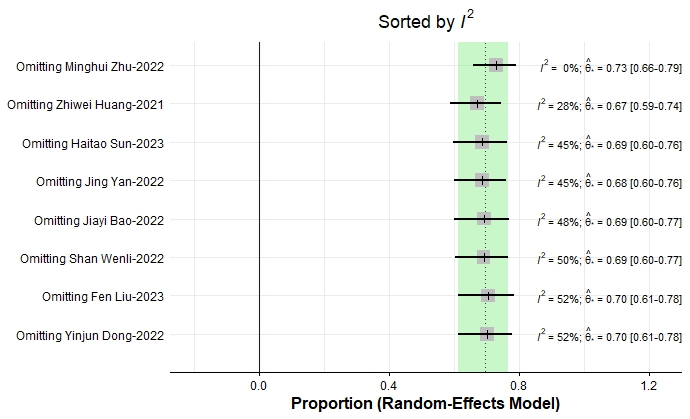

**Figure 1(Supp).** Heterogeneity of studies (A) Sensitivity – Training cohort, (B) Specificity – Training cohort, (C) Specificity – Validation cohort
