## Supplementary material - Keywords of Radiomic-ki67 for "Diagnostic Performance of Radiomics in Prediction of Ki-67 Index Status in Non-small Cell Lung Cancer: A Systematic Review and Meta-Analysis"

“Radiomic Keywords”

MESH Terms:

.

Emtree Terms:

radiomics

SYNONYMS:

Radiomic*

“lung Keywords”

MESH Terms:

.

Emtree Terms:

.

SYNONYMS:

Lung

pulmonary

“Ki-67 Keywords”

MESH Terms:

.

Emtree Terms:

.

SYNONYMS:

Ki-67

Ki67

MIB-1

MIB1
