## Supplementary material - RQS_table_1 for "Diagnostic Performance of Radiomics in Prediction of Ki-67 Index Status in Non-small Cell Lung Cancer: A Systematic Review and Meta-Analysis"

**Table 1S.** Basic adherence rate according to the six key domains

|  | **Basic adherence rate** |
| --- | --- |
| Total 16 items | 33.3% |
| Domain 1: Protocol quality and stability in image and segmentation | 28% |
| Protocol quality | 10 (100%) |
| Test-retest | 0 (0%) |
| Phantom study | 0 (0%) |
| Multiple segmentation | 4 (40%) |
| Domain 2: Feature selection and validation | 61.3% |
| Feature reduction or adjustment of multiple testing | 10 (100%) |
| Validation | 9 (90%) |
| Domain 3: Biologic/clinical validation and utility | 46.7% |
| Multivariate analysis with non-radiomics features | 8 (80%) |
| Biologic correlates | 6 (60%) |
| Comparison to ‘gold standard’ | 0 (0%) |
| Potential clinical utility | 7 (70%) |
| Domain 4: Model performance index | 58% |
| Discrimination statistics | 10 (100%) |
| Calibration statistics | 5 (50%) |
| Cut-off analysis | 3 (30%) |
| Domain 5: High level of evidence | 0% |
| Prospective study | 0 (0%) |
| Cost-effective analysis | 0 (0%) |
| Domain 6: Open science and data | 0 (0%) |
