## Supplementary material - RQS_table_2 for "Diagnostic Performance of Radiomics in Prediction of Ki-67 Index Status in Non-small Cell Lung Cancer: A Systematic Review and Meta-Analysis"

**Table 2S.** Details of quality assessment by Radiomics Quality Score (RQS) of all included studies

| First Author | Image protocol quality | Multiple segmentations | Phantom study on all scanners | Imaging at multiple time points | Feature reduction or adjustment for multiple testing | Multivariable analysis with non-radiomics features | Detect and discuss biological correlates | Cut-off analyses | Discrimination statistics | Calibration statistics | Prospective study registered in a trial database | Validation | Comparison to gold standard | Potential clinical utility | Cost-effectiveness analysis | Open science and data | Total |
| --- | --- | --- | --- | --- | --- | --- | --- | --- | --- | --- | --- | --- | --- | --- | --- | --- | --- |
| Bao 2022 | 1 | 0 | 0 | 0 | 3 | 1 | 1 | 0 | 2 | 2 | 0 | 2 | 0 | 2 | 0 | 0 | 14 (38.89%) |
| Wenli 2022 | 1 | 1 | 0 | 0 | 3 | 1 | 0 | 0 | 2 | 0 | 0 | 2 | 0 | 2 | 0 | 0 | 12 (33.33%) |
| Huang 2021 | 1 | 0 | 0 | 0 | 3 | 1 | 1 | 1 | 2 | 0 | 0 | 2 | 0 | 0 | 0 | 0 | 11 (30.55%) |
| Yan 2022 | 1 | 0 | 0 | 0 | 3 | 1 | 1 | 0 | 1 | 0 | 0 | 2 | 0 | 2 | 0 | 0 | 11 (30.55%) |
| Zhu 2022 | 1 | 0 | 0 | 0 | 3 | 1 | 1 | 0 | 2 | 0 | 0 | 2 | 0 | 0 | 0 | 0 | 10 (27.78%) |
| Sun 2023 | 1 | 1 | 0 | 0 | 3 | 0 | 0 | 0 | 2 | 2 | 0 | 2 | 0 | 2 | 0 | 0 | 13 (36.11%) |
| Gu 2019 | 1 | 0 | 0 | 0 | 3 | 0 | 0 | 0 | 2 | 0 | 0 | -5 | 0 | 0 | 0 | 0 | 1 (2.78%) |
| Dong 2023 | 1 | 1 | 0 | 0 | 3 | 1 | 1 | 1 | 2 | 2 | 0 | 5 | 0 | 2 | 0 | 0 | 19 (52.78%) |
| Liu 2023 | 1 | 1 | 0 | 0 | 3 | 1 | 0 | 1 | 2 | 1 | 0 | 5 | 0 | 2 | 0 | 0 | 17 (47.22%) |
| Yao 2022 | 1 | 0 | 0 | 0 | 3 | 1 | 1 | 0 | 1 | 1 | 0 | 2 | 0 | 2 | 0 | 0 | 12 (33.33%) |
