## Supplementary material - Search Queries of Radiomic-ki67 for "Diagnostic Performance of Radiomics in Prediction of Ki-67 Index Status in Non-small Cell Lung Cancer: A Systematic Review and Meta-Analysis"

Pubmed Search Query

Time of search : 16 November 2023
Results : 22

(Radiomic*[tiab]) AND (Lung[tiab] OR pulmonary[tiab]) AND (Ki-67[tiab] OR Ki67[tiab] OR MIB-1[tiab] OR MIB1[tiab])

Embase Search Query

Time of search : 16 November 2023
Results : 27

(‘Radiomics’/exp OR ‘Radiomic*’:ab,ti) AND (‘Lung’:ab,ti OR ‘pulmonary’:ab,ti) AND (‘Ki-67’:ab,ti OR ‘Ki67’:ab,ti OR ‘MIB-1’:ab,ti OR ‘MIB1’:ab,ti)

Scopus Search Query

Time of search : 16 November 2023
Results : 23

(TITLE-ABS (“Radiomic*”)) AND (TITLE-ABS (“Lung” OR “pulmonary”)) AND (TITLE-ABS (“Ki-67” OR “Ki67” OR “MIB-1” OR “MIB1”))

Web of science Search Query

Time of search : 16 November 2023
Results : 25

(TS= (“Radiomic*”)) AND (TS= (“Lung” OR “pulmonary”)) AND (TS= (“Ki-67” OR “Ki67” OR “MIB-1” OR “MIB1”))
